# What is next for costing WASH in healthcare facilities? Applying evidence for policy and practice

**DOI:** 10.1101/2024.08.14.24311992

**Authors:** Lucy K. Tantum, Ryan Cronk, Narathius Asingwire, Prakash Bohara, Laxman Kharal Chettry, Mitsuaki Hirai, Sena Kpodzro, Tariro Mavi, Joshua D. Miller, Carrie Ripkey, Sarbesh Sharma, Victoria Trinies, Darcy Anderson

## Abstract

Efforts to improve and sustain water, sanitation, hygiene (WASH), waste management, and cleaning services in healthcare facilities in low- and middle-income countries are constrained by limited funding. Assessments of the costs of delivering WASH services are critical for guiding financial planning and investment, but many countries lack costed plans for WASH in healthcare facilities. A 2023 UNC Water and Health Conference workshop explored how policymakers and practitioners collect and use cost data and identify strategies for overcoming cost barriers. Presenters shared case studies that showcased the utility of cost data for creating national costed roadmaps, identifying and addressing budgetary shortfalls, and planning WASH improvements in Nepal, Uganda, and Zimbabwe. In discussions, workshop participants described leveraging collaborations with multiple government entities and non-governmental organizations (NGOs) to collect cost data. Participants also reported using cost data to plan programs and advocate for additional WASH funding. Strategies to coordinate costing approaches across stakeholder groups and standardize data collection tools will enhance the efficiency and effectiveness of planning and budgeting for WASH in healthcare facilities.

**Highlights:**

1. Local and national governments and NGOs routinely collect and use cost data to inform decisions about WASH services in healthcare facilities
2. WASH responsibilities are divided across multiple institutions, necessitating coordination for cost data collection
3. Governments have used costing data to create national costed roadmaps and budgets
4. Costing data support program planning, monitoring, and advocacy

## Introduction

Reliable access to water, sanitation, hygiene (WASH), waste management, and cleaning is essential for safe and high-quality healthcare delivery [1,2]. Improving and sustaining WASH in healthcare facilities is a global priority, particularly in low- and middle-income countries where many healthcare facilities lack adequate WASH services [3,4]. Achieving basic water, sanitation, hygiene, and waste management services in least-developed countries is estimated to cost US$ 2.9-4.8 billion as a one-time investment, with an additional US$ 0.39-0.60 per capita per year to sustain those services [5]. Additional investments will be needed to achieve and sustain environmental cleaning services and strengthen the energy infrastructure necessary to operate many WASH services (e.g., electrically pumped boreholes and autoclaves). These costs are difficult to estimate over the lifecycle of WASH services [6].

Investments in infection control and WASH in healthcare facilities can yield positive returns, as WASH improvements can avert infections and reduce healthcare costs [7]. While government spending on healthcare has increased in recent years, investments in WASH in healthcare facilities have lagged behind [8]. In 2022, 3% of surveyed low- and middle-income countries reported having adequate financial and human resources to implement policies to improve WASH in healthcare facilities [9]. Financial investment thus remains a substantial barrier to sustainable improvement.

Costing and budgeting inform investments to achieve and sustain WASH in healthcare facilities. Costing involves estimating the expenses required to deliver goods and services over a defined timespan. For example, one approach may be to enumerate the activities and inputs required to achieve a basic waste management service and then calculate the overall costs based on their quantities and unit prices. Budgeting involves determining how much money will be spent over time, the funding source, and what specific line items will be purchased. Costing can help determine the total service delivery costs and ensure sufficient budgets. Budgeting can, however, be done without costing when the total amount of funding is pre-determined, and stakeholders must create a plan to spend it [10,11].

In 2019, the World Health Organization (WHO) and United Nations Children’s Fund (UNICEF) released *Eight Practical Steps*, a guideline document that outlines eight recommended steps for achieving universal WASH in healthcare facilities. Step two in this guideline encourages countries to develop WASH policies with costed roadmaps [12]. Costed roadmaps include well-defined targets, mechanisms for tracking progress, and estimates of the costs required to achieve and maintain goals [12]. To date, 24% of countries have developed and approved a national policy and costed roadmap for WASH in healthcare facilities [9].

Barriers to creating costed roadmaps include limited evidence on the costs of WASH services, tools for collecting data, and guidance on applying data for policy and practice [6]. Several studies have estimated the costs of WASH in healthcare facilities at regional, national, and international levels and highlighted the need for additional investment [5,6,13,14]; however, large gaps remain. Most studies are conducted in low-income countries and measure progress using the “basic” service levels developed as targets for the Sustainable Development Goals [15]. Middle-income countries are underrepresented, as are more advanced service levels beyond “basic,” which may be required for specific settings or facility types. Toolkits are available to support practitioners in costing WASH in healthcare facilities and address these data gaps [16,17]. Importantly, no studies describe how these tools and the resulting data they generate can be applied to policy and practice.

We hosted a workshop on evidence, policy, and practice for costing WASH in healthcare facilities at the University of North Carolina (UNC) Water and Health Conference on October 26, 2023. The Water Institute at UNC, the United States Centers for Disease Control and Prevention (CDC), the CDC Foundation, the Nepal Ministry of Health and Population, Terres de hommes Nepal, Helvetas, and the Swiss Water and Sanitation Consortium convened the workshop. Objectives were to 1) document available tools, approaches, and practices for costing WASH in healthcare facilities; 2) describe how cost data can be applied to inform policy and practice; and 3) evaluate barriers experienced by policymakers and practitioners for applying cost data for evidence-based decision-making. This paper synthesizes the presentations and breakout discussions that occurred during the event.

## Methods and event description

During the conference workshop, hosting organizations presented background information on costing WASH in healthcare facilities and shared case studies of successful costing projects in three countries: (1) analyzing budgets and financing systems for WASH in healthcare facilities in Uganda (CDC, CDC Foundation, Uganda Socio-Economic Data Centre), (2) leveraging cost data to develop municipal WASH operations and maintenance policies and inform national costed roadmaps (Nepal Ministry of Health and Population, Terre des hommes Nepal, Helvetas, Swiss Water and Sanitation Consortium), and (3) developing a costing tool for programs using the Water and Sanitation for Health Facility Improvement Tool (WASH FIT) (UNICEF Zimbabwe).

Breakout discussions followed presentations. Participants were self-selected into two breakout groups to discuss approaches to collecting and using cost data for WASH in healthcare facilities, barriers to obtaining and applying cost data, and strategies to overcome barriers. A facilitator-guided discussion for each group (See Supplementary Information 1 for discussion questions). At the end of the workshop, participants reconvened to share and synthesize findings from breakout discussions, identifying new learnings and common themes that emerged across the groups. Four note-takers (two per breakout group) recorded the conversation throughout the event. All note-takers transcribed their notes following the event, and two researchers triangulated findings across note-takers to ensure all relevant information was accurately captured. This manuscript summarizes country case studies and synthesizes breakout group discussions.

## Results

### Country case studies

#### Case study 1: District-level budgeting and financing analysis in Uganda

The Uganda case study presented the results of a project to understand the national and district-level systems in place to support financing and budgeting for WASH in healthcare facilities. Between 2022 and 2023, the CDC and the CDC Foundation supported the Socio-Economic Data Centre in conducting a review of existing materials (e.g., policies, strategic plans, and budget documents) and primary data collection via interviews with 80 key informants at the national level and in Kabarole and Lira Districts.

Thematic analysis of primary and secondary data revealed two major sources of financing for healthcare facilities in Uganda: *on-budget financing*, appropriated by the National Parliament and passed through the Central Government Treasury, and *off-budget financing*, which included resources provided directly by donors. A review of district and facility-level budgets from 2019-2023 indicated that off-budget financing was the primary source of WASH in healthcare facilities. Additionally, funding streams were not well coordinated across national and sub-national levels, with several streams contributing to the same types of activities (e.g., construction of new hygiene infrastructure) and few streams contributing to other areas such as construction of new sanitation infrastructure or ongoing operations and maintenance, the latter major challenges to infrastructure sustainability.

Critical strategic planning documents, including the National Development Plan, determine budget priorities at the highest level and inform Ministry-level strategic planning documents such as the Ministry of Health Strategic Plan [18]. Based on the priorities outlined in these documents, the overall amount of funds allocated across budget lines is constrained by the resource envelope established by the Medium-Term Expenditure Framework [18]. These strategic planning documents minimally mention institutional WASH so that few funds can be allocated to WASH in healthcare facilities [19,20]. Budget development processes are, however, highly consultative, providing an opportunity to advocate for greater inclusion of WASH in the future [21].

#### Case study 2: Costed roadmap development in Nepal

The Nepal case study described the process used by the Management Division under the Ministry of Health and Population of Nepal, in collaboration with Terre des hommes, to enhance WASH in healthcare facilities across the country through policies and costed roadmaps. This collaboration was pivotal in developing a data-driven approach to formulating WASH policies and costed roadmaps, ensuring that every healthcare facility in Nepal has access to necessary WASH services. Recognizing a need for detailed financial planning, the Ministry of Health and Population prioritized the development of costed roadmaps for WASH in healthcare facilities, aiming to place Nepal among the global frontrunners to achieve universal WASH in healthcare settings.

In 2022, Terre des hommes conducted a comprehensive costing exercise in Thakurbaba Municipality, providing essential data on capital and annual operation and maintenance costs for WASH services. The data generated from this exercise shaped the Ministry of Health and Population’s understanding of actual costs and demonstrated how local data can inform national policy. The insights gained from Terre des homme’s study were integrated into Nepal’s National Roadmap for WASH in Healthcare Facilities. This integration informed budget allocations and enriched policy guidelines, particularly in operations and maintenance, resulting in effective, evidence-based policymaking to drive sustainable outcomes.

In 2023, the Ministry of Health and Population submitted the final draft of the National Roadmap to relevant higher-level government authorities for approval. The Ministry aimed to implement the roadmap, monitor its impact, and make adaptations based on emergent data and insights in continued collaboration with development partners, including Terre des hommes, and with support from the Swiss Water and Sanitation Consortium. The costed roadmap is envisioned as a financial blueprint and a multifaceted tool, strategically aligning stakeholders towards shared goals.

#### Case study 3: WASH FIT costing tool in Zimbabwe

The Zimbabwe case study presented the outcomes of a project to develop a costing tool to inform budgeting and resource mobilization to improve WASH services in healthcare facilities. In 2021, the Ministry of Health and Child Care led the Water and Sanitation for Health Facility Improvement Tool (WASH FIT) rollout in 100 healthcare facilities, following an initial pilot in 2020 during COVID-19 [22,23]UNICEF, the CDC, and USAID’s Bureau for Humanitarian Assistance provided financial and technical assistance for the WASH FIT assessment and financed WASH upgrades in target healthcare facilities from 2021 to 2023.

While the WASH FIT assessment allowed healthcare facilities and the Government of Zimbabwe to collect information on the status of available WASH services, little was known about the costs of improving WASH services in healthcare facilities. Developing a tool that would provide cost estimates was essential to understand the resources needed to achieve at least basic WASH services in healthcare facilities. Cost estimates enabled healthcare facility staff to understand facility-level resource needs. The assessment also enabled the Government to estimate a national budget requirement to mobilize resources from the Government and development partners.

Based on the 2021 WASH FIT assessment results and identified priority actions, detailed costing was conducted to enable the WASH service upgrades. Based on the detailed costing information generated from the actual improvement of WASH services and the WASH FIT assessment results, an Excel-based WASH FIT Costing Tool was developed in 2022. The WASH FIT Costing Tool was digitized and incorporated into the Ministry of Health and Child Care’s information systems in 2023.

When applied, the WASH FIT Costing Tool presents a total budget requirement per healthcare facility by estimating costs for capital (infrastructure upgrades), software (training), and operational requirements (supplies and minor repairs). The WASH FIT Costing Tool estimated a preliminary national budget requirement of $116 million to upgrade WASH services in all healthcare facilities. This informed the Government’s budgeting decisions, WASH in healthcare facility program planning, and resource mobilization by the Government and its partners. While the accuracy and reliability of estimated budget requirements can be iteratively improved as additional cost data are collected, the Government of Zimbabwe and its partners gained insights into the scale of financial resources needed to provide WASH services that underpin quality care and infection prevention and control in healthcare facilities.

### Breakout group discussions

Approximately 40 participants participated in breakout discussions. Participants were affiliated with government institutions, local and international NGOs, and academic centers in sub-Saharan Africa, the Middle East, South and Southeast Asia, Europe, and North America. In breakout groups, workshop participants discussed their experiences with costing WASH in healthcare facilities, including strategies to collect cost data, overcome barriers to costing, and apply cost data in practice. Some participants shared experiences with generating and using cost data in policy contexts, such as in regional government, national government, and as an advocacy strategy. Others used costing in a programmatic context, such as NGOs implementing WASH improvement programs. Key takeaways from these discussions are provided below.

#### Government and non-governmental stakeholder groups collect and use cost data

Approaches to costing WASH in healthcare facilities varied across country contexts, depending on the availability of existing tools, data sources, and stakeholder involvement. For most breakout group participants, costing was more common and straightforward for program start-up activities (e.g., infrastructure construction, initial training), as many organizations had databases that provided costs of commonly used supplies or could contact local contractors for this information. Costing was more challenging and less frequent for long-term expenses related to operations and maintenance, as factors such as infrastructure repair needs and changes in demand for services were difficult to predict but had substantial cost implications.

In many countries, robust data on the costs of WASH in healthcare facilities were unavailable, leading stakeholders to extrapolate the costs of WASH services based on those reported in other settings. For example, a citywide sanitation program in Indonesia developed a national costing tool that aggregated data from global estimates, national sources, and city-level sources. In Palestine, WASH costing analysis was done through schools, focusing on distinguishing between preventive maintenance and repair costs. Costing sessions were initiated by meetings to review financial roles and assess funds for priority setting. In Kenya, WASH costing was prompted by evaluating different school WASH programs, such as hand hygiene stations. Cost data extraction was complex, as it was not routinely collected and was sensitive to procurement issues.

The extent to which governments were engaged in cost data collection varied. Nepal’s government regularly collected cost data for its WASH budgets and development roadmaps. In contrast, in Ghana, there were fewer systematic data collection efforts, so many organizations rely on NGOs’ ad hoc data collection efforts. Additionally, participants noted discrepancies over which agencies should be responsible for collecting cost data, such as tensions between the Ministry of Health and the Ministry of Water over responsibilities for data collection.

#### Cost data are used to plan programs and advocate for funding

Breakout groups identified two applications for cost data: program management and advocacy. For program planning and management, cost data were used to understand program expenses over time and create budgets sufficient to sustain program activities over the intended program lifespan–though predicting long-term costs was challenging, and budgets were sometimes insufficient. Cost data were also used to monitor and evaluate programs’ progress and impact and identify areas for improvement. In some contexts, cost data were used to demonstrate the benefits of preventive maintenance over repair and estimate the potential savings and efficiency gains to raise awareness and mobilize resources from the community and donors. Lastly, cost data were used to compare the effectiveness and sustainability of different WASH interventions to inform the design and implementation of future programs.

For policy and advocacy, cost data were primarily used by local and national governments to evaluate the feasibility of WASH upgrades and develop practical plans for scaling up and maintaining high-quality services. Several breakout group attendees mentioned using cost data for nationally costed roadmaps, as described in guidelines from the WHO and UNICEF (WHO/UNICEF 2019). Cost data were used for advocacy by NGOs to demonstrate budgetary shortfalls and advocate for funding from the government. In some cases, governments also used data for advocacy. For example, local governments advocated to national governments for more funding or governments advocated to external development partners. Results demonstrating potential cost savings and health improvements were compelling evidence to garner greater interest and investment in WASH upgrades and upkeep. However, the utility of these findings varied depending on political power dynamics.

#### Data collection requires standardization of costing tools and collaboration among stakeholders

Barriers to cost data collection included technical difficulties related to variability and uncertainty of cost estimates due to market fluctuations and dynamic exchange rates. Capturing lifecycle costs was difficult due to changes in population size, shifting needs, and uncertain repair costs. Indirect costs of WASH, such as supervision and management, were important but difficult to quantify. Participants also reported barriers to data collection related to stakeholder roles and relationships. The lack of transparency and accountability in the procurement and budgeting processes made it difficult to estimate actual costs. The limited involvement of key actors also hindered the costing process. In some cases, participants noted that cost data could be sensitive and that some stakeholders may be unwilling to share information.

Developing and adapting standardized, user-friendly tools was one strategy to address challenges with stakeholder coordination and data inconsistency and scarcity. Integrating WASH costing tools into existing data collection and management systems helped streamline and routinize data collection to address data scarcity issues.

#### Coordination and transparency during the costing process can improve buy-in and data usage

Participants reported barriers to data use, including limited coordination between stakeholders for generating and using data; inconsistencies in data formats, WASH technologies, or activities; and data literacy among decision-makers and community leaders. Engaging community advisory groups was identified as a strategy to help determine local needs and priorities when calculating WASH budgets. In cases where data were collected and used by different types of stakeholders, the mismatch between donor funding cycles and project cycles complicated budgeting efforts. Furthermore, some stakeholders noted that governments did not prioritize WASH even when data were available to make an investment case.

Co-designing data collection systems successfully addressed data use challenges, strengthened data transparency, and built trust. In Indonesia, a WASH costing tool was integrated with a WASH FIT program using a participatory design process, enhancing stakeholder buy-in. Participants emphasized the importance of leveraging support from multiple partners and demonstrating the cross-sectoral impacts of investing in WASH (e.g., improvements to child health and development) to fund data collection. Other suggested strategies included using photos, testimonies, and personal narratives as tools for advocacy related to WASH financing and competitions focused on WASH milestones (e.g., the first community to have improved water sources for all healthcare facilities) to enhance political will.

## Discussion

During this workshop, case studies and breakout discussions on costing WASH in healthcare facilities revealed strategies to improve WASH policy and programming effectiveness by collecting and applying cost data. Case studies demonstrated the importance of understanding WASH financial processes in healthcare facilities to integrate costing into existing systems. Cost data are helpful in planning and budgeting processes for WASH in healthcare facilities at the facility, regional, and national levels. Additionally, costing plays a role in the selection and evaluation of WASH interventions and in efforts to advocate for increased funding. Findings highlight the importance of fostering collaboration across stakeholder groups and standardizing data collection methods to improve cost data availability, quality, and utility.

Coordination among the stakeholders involved in costing WASH in healthcare facilities posed a barrier to large-scale collection and usage of cost data. Workshop participants reported that costing tools and approaches were not standardized across institutions, and data collection lacked transparency. As a result, it was difficult to describe the lifecycle costs of a WASH program fully. Despite these challenges, participants succeeded in introducing standardized costing tools, coordinating data collection across stakeholder groups, and co-designing data collection systems with end users. Effective collection and application of cost data requires a complete understanding of the stakeholders involved in WASH service delivery [10]. Stakeholder mapping strategies have previously been used to understand roles for delivering WASH services in community settings [24,25]. These methods could be applied to healthcare facility contexts. Furthermore, stakeholders can use standard tools developed for costing WASH in healthcare facilities for more efficient and complete data collection [16].

In both policy and program contexts, pairing cost data with health information- and non-health impacts of interventions effectively raised awareness of the importance of WASH in healthcare facilities. They built an investment case for specific interventions. Concurrent cost and impact data collection could support evidence-based decision-making around investments for WASH in healthcare facilities. Prior research has applied cost-benefit or cost-effectiveness evaluation for healthcare waste management and hand hygiene interventions in low-resource contexts [26–28]. The World Health Organization’s 2023 infection prevention and control strategy describes the importance of building an investment case for WASH in healthcare facilities, highlighting an opportunity to integrate WASH costing with broader health system financing strategies. More research is needed to characterize and measure the potential benefits of WASH in healthcare facilities, such as physical and mental health, healthcare worker wellbeing, patient satisfaction, and health system resilience [6]. Where data on these benefits are lacking, stakeholders can consider presenting intervention narratives and case studies alongside cost data to demonstrate the impact of investment in WASH in HCFs.

## Conclusions

Understanding the costs associated with delivering WASH services in healthcare facilities can guide planning and decision-making to improve and sustain WASH services in healthcare facilities. Practitioners and researchers should work to strengthen and standardize cost data quality, scale up costing approaches to understand lifecycle costs better and explore strategies to use cost data to advocate for policy change. Collecting and using cost data is essential in accelerating improvement toward universal safe WASH.

## Data Availability

All data produced in the present work are contained in the manuscript.

## Acknowledgments

We thank the workshop hosts, presenters, and participants for contributing their experiences and viewpoints on costing WASH in healthcare facilities. We thank Cindy Kushner for providing feedback on the draft manuscript.

## Disclaimer

The perspectives expressed are those of the authors and do not necessarily reflect the views of their institutions, Governments, UNICEF, and the United Nations. The findings and conclusions of this report are those of the authors and do not necessarily represent the official position of the Centers for Disease Control and Prevention (CDC).

