## Supplementary Information 1 for "What is next for costing WASH in healthcare facilities? Applying evidence for policy and practice"

**Supplementary information 1: Breakout discussion guide**

What are your approaches for costing WASH in healthcare facilities in your organization or country?

- Does your organization collect costs data? If so, what data? How is it collected?
- Does your organization receive costs data from others? If so, what type of data?
- Do you participate in any monitoring systems that collect indicators on costs as part of routine monitoring? What indicators do you track?
- What is your biggest unmet need for costs data? Why is this need unmet?

How does your organization or country use costs data for WASH in healthcare facilities policy and practice?

- What are the biggest challenges related to financing WASH in healthcare facilities in your context? Can data help solve those challenges? How?
- Is your organization or country currently using any available costs data for decision making? If yes, how? If no, why not?

What are the biggest barriers to collecting costs data and applying it for policy and practice?

- What kinds of barriers do you experience related to data availability? Data quality? Data collection? Capacity for understanding and applying costs data?
- If you are a data collector, what do you wish policy makers knew?
- If you are a data user, what do you wish data collectors knew?

What strategies have you employed to overcome these barriers?

- Tell us about a success story from your context. What is one example of how you used evidence or data to solve a challenge related to WASH in healthcare facilities financing?
- How do you make decisions about budgeting and financing when you lack adequate costing data?
- What are some ways to improve collaboration between researchers and policy makers?
